# Rare variant aggregation highlights rare disease genes associated with brain volume variation

**DOI:** 10.1101/2024.09.26.24314187

**Authors:** Douglas P. Wightman, Bernardo A.P.C. Maciel, Rachel M. Brouwer, Martijn P. van den Heuvel, Danielle Posthuma

**Affiliations:** Department of Complex Trait Genetics, Center for Neurogenomics and Cognitive Research, Amsterdam Neuroscience, Vrije Universiteit Amsterdam, Amsterdam, The Netherlands; Department of Child and Youth Psychiatry, Emma Children’s Hospital, Section Complex Trait Genetics, Amsterdam Neuroscience, Amsterdam UMC, 1081 HV Amsterdam, The Netherlands

## Abstract

Variation in brain volume is associated with common and rare disorders. Investigating the genetics of brain volume may highlight overlap between diseases and biological mechanisms that explain disease symptoms. Previous studies examining genetic variants associated with brain volume have largely focused on common variants, with rare variant studies not primarily focusing on brain volume phenotypes or focusing on large structural variants. In this study, we aggregated rare variants within genes and associated genes with 44 brain volume phenotypes in the UK Biobank (N=40,374). We found convergence between genes within the same biological pathway and convergence between common and rare variants within the same gene. Seven of the genes associated with total or regional brain volume measures were also linked with rare brain disorders in previous literature. We successfully showed that rare variants in genes linked to rare brain disorders were also associated with sub-clinical brain volume variation.

## Main text

Variation in brain volume is associated with common^1,2^ and rare disorders^3^. Investigating the genetics of brain volume may highlight overlap between diseases and biological mechanisms that explain disease symptoms. Previous studies examining genetic variants associated with brain volume have largely focused on common variants^4–8^, with rare variant studies not primarily focusing on brain volume phenotypes^9–11^ or focusing on large structural variants^12,13^. In this study, we aggregated rare variants within genes and associated genes with 44 brain volume phenotypes in the UK Biobank (N=40,374). We found convergence between genes within the same biological pathway and convergence between common and rare variants within the same gene. Seven of the genes associated with total or regional brain volume measures were also linked with rare brain disorders in previous literature. We successfully showed that rare variants in genes linked to rare brain disorders were also associated with sub-clinical brain volume variation.

We aggregated rare (MAF<0.01) missense and predicted loss-of-function (pLOF) variants across 18,613 genes to assess gene associations with total brain volume (40,374 individuals) and 43 cortical or subcortical volume measures (36,709 individuals). A full list of the tested brain volume measures is available in **Supplementary Table 1**. We used a gene-based unified test to identify genes associated with these phenotypes (**Methods**). These results were compared to a null model created by repeating the analyses using only synonymous variants (not expected to impacted the phenotype). We used the results from the gene association analyses to highlight genetic overlap between brain volume variation and rare brain disorders, to prioritise genes relevant for brain volume differences, and to identify gene-sets and tissues relevant to brain volume variation (**Figure 1**).

**Figure 1:**
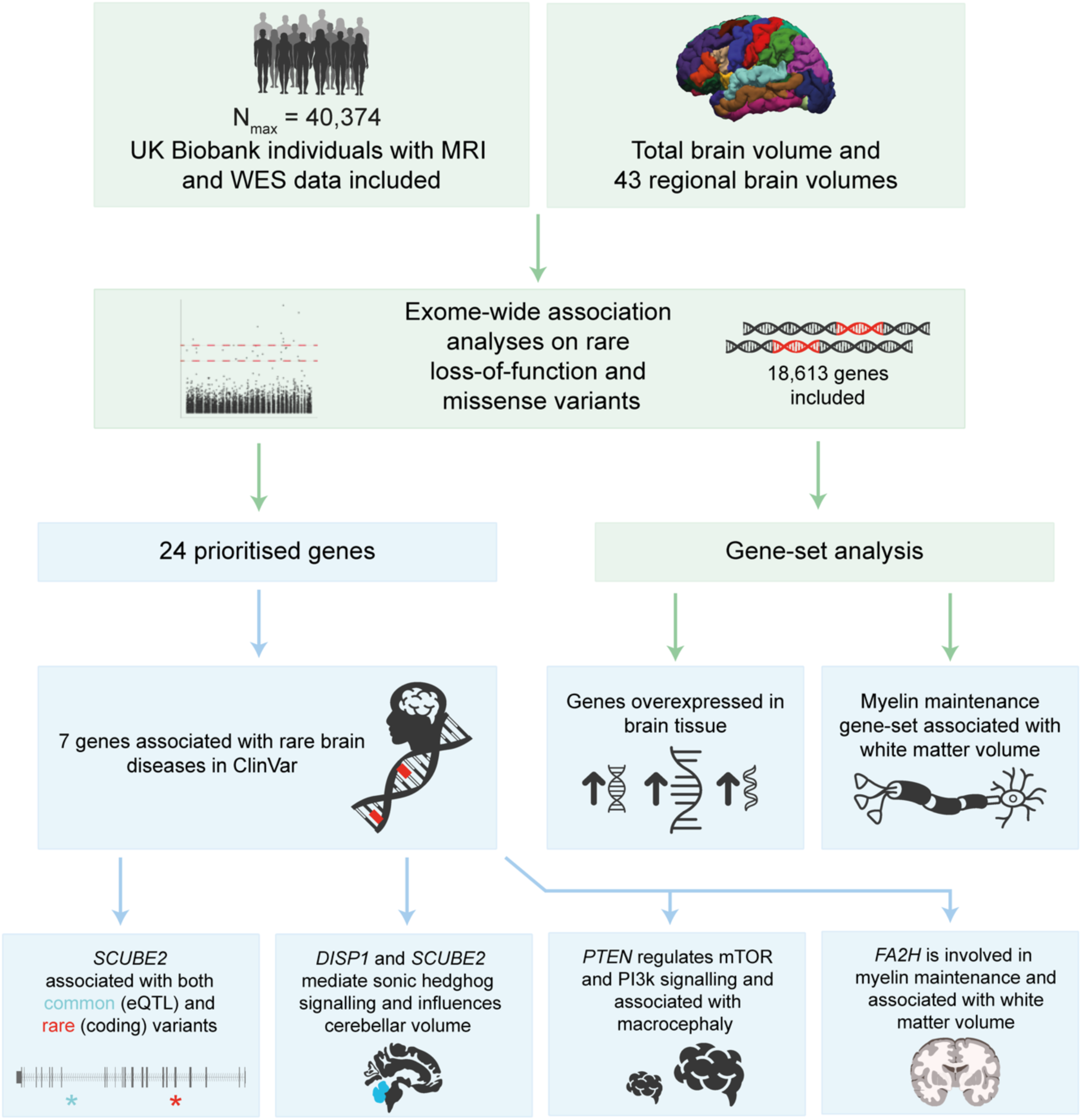
A visual summary of the methods and results of this study shows that we identified 24 genes associated with brain volume variation through rare variants, 7 of which were associated with rare brain diseases in a database of clinically relevant genetic variation (ClinVar). WES represents whole exome sequencing. Gene-set analysis results are reported in **Supplementary Note**.

## Gene results

We used the P-value of the unified test (GENE_P) to prioritise genes at two levels; genome wide significant (GWS) and study wide significant (SWS). The GWS genes were defined as genes with a P-value <0.05 after Bonferroni correction within each phenotype (0.05/18613 genes= 2.69×10^-6^) and the SWS genes were defined as genes with a P-value <0.05 after Bonferroni correction across all phenotypes (0.05/(18613 genes*44 phenotypes)= 6.11×10^-8^). We identified 26 GWS gene-phenotype associations (24 unique genes) with 8 of those also being SWS (**Table 1**; **Figure 2**; **Supplementary Table 2**). In our negative control (synonymous variants only), there was only 1 SWS gene and 10 GWS genes. None of these synonymous associated genes overlapped with the pLOF+missense associated genes. This suggests that the false positive rate was relatively well calibrated.

**Table 1:** The 26 gene-phenotype associations from the pLOF+missense variant analyses show 7 genes associated with differences in a brain volume that have been reported as relevant for a brain disease in ClinVar. Bankssts represents Banks of the Superior Temporal Sulcus and Rostralanterior represents rostral anterior cingulate cortex. SWS represents whether that association is study wide significant (P<6.11×10^-8^), cMAC represent the cumulative minor allele count of the variants that mapped to that gene in the most significant analysis of that gene, # of variants represents the number of variants that mapped to that gene in the most significant analysis of that gene.

| Phenotype | Gene name | SWS | P-value | cMAC | # of variants | Brain ClinVar annotation |
| --- | --- | --- | --- | --- | --- | --- |
| Cortical grey matter | *FA2H* | 1 | $3.12 \times 10^{-12}$ | 76 | 28 | Cerebellar atrophy |
| White matter | *FA2H* | 1 | $1.16 \times 10^{-8}$ | 76 | 28 | Cerebellar atrophy |
| Cerebellum | *DISP1* | 1 | $1.36 \times 10^{-8}$ | 1273 | 223 | Microform holoprosencephaly |
| Cerebellum | *SCUBE2* | 0 | $6.73 \times 10^{-8}$ | 770 | 195 | Cerebral arteriovenous malformation |
| Brainstem | *NDUFA2* | 0 | $1.94 \times 10^{-7}$ | 6 | 5 | Cystic Leukoencephalopathy |
| Total brain volume | *PTEN* | 0 | $2.61 \times 10^{-7}$ | 6 | 6 | Macrocephaly |
| Caudate | *GPATCH11* | 0 | $3.54 \times 10^{-7}$ | 18 | 11 | Early onset and severe retinal dystrophy with neurological impairment and facial dysmorphism |
| Bankssts | *KMT5B* | 0 | $8.14 \times 10^{-7}$ | 847 | 97 | Neural tube defect |
| White matter | *RPS21* | 1 | $1.94 \times 10^{-11}$ | 1 | 1 | - |
| Cortical grey matter | *POLD4* | 1 | $6.14 \times 10^{-10}$ | 1 | 1 | - |
| insula | *PKD1* | 1 | $3.00 \times 10^{-9}$ | 12240 | 1373 | - |
| White matter | *ZFR2* | 1 | $1.96 \times 10^{-8}$ | 2191 | 277 | - |
| Pars opercularis | *ADAMTS18* | 1 | $2.54 \times 10^{-8}$ | 228 | 87 | - |
| White matter | *DOC2A* | 0 | $6.00 \times 10^{-8}$ | 137 | 29 | - |
| pericalcarine | *TNS3* | 0 | $6.04 \times 10^{-8}$ | 389 | 67 | - |
| Total brain volume | *EVI2A* | 0 | $1.04 \times 10^{-7}$ | 1272 | 50 | - |
| White matter | *CENPO* | 0 | $1.26 \times 10^{-7}$ | 707 | 67 | - |
| White matter | *C17orf100* | 0 | $1.27 \times 10^{-7}$ | 2 | 2 | - |
| White matter | *ZBTB48* | 0 | $2.07 \times 10^{-7}$ | 4 | 4 | - |
| Total brain volume | *SPSB3* | 0 | $3.27 \times 10^{-7}$ | 1328 | 85 | - |
| Hippocampus | *ADCY6* | 0 | $5.21 \times 10^{-7}$ | 122 | 78 | - |
| Pars opercularis | *CCM2L* | 0 | $9.46 \times 10^{-7}$ | 755 | 127 | - |
| Brainstem | *PRUNE2* | 0 | $1.36 \times 10^{-6}$ | 4253 | 488 | - |
| Rostral anterior cingulate | *SPMIP1* | 0 | $1.77 \times 10^{-6}$ | 3 | 3 | - |
| precuneus | *RPUSD1* | 0 | $1.78 \times 10^{-6}$ | 691 | 112 | - |
| Cortical grey matter | *RPS21* | 0 | $2.38 \times 10^{-6}$ | 1 | 1 | - |

**Figure 2:**
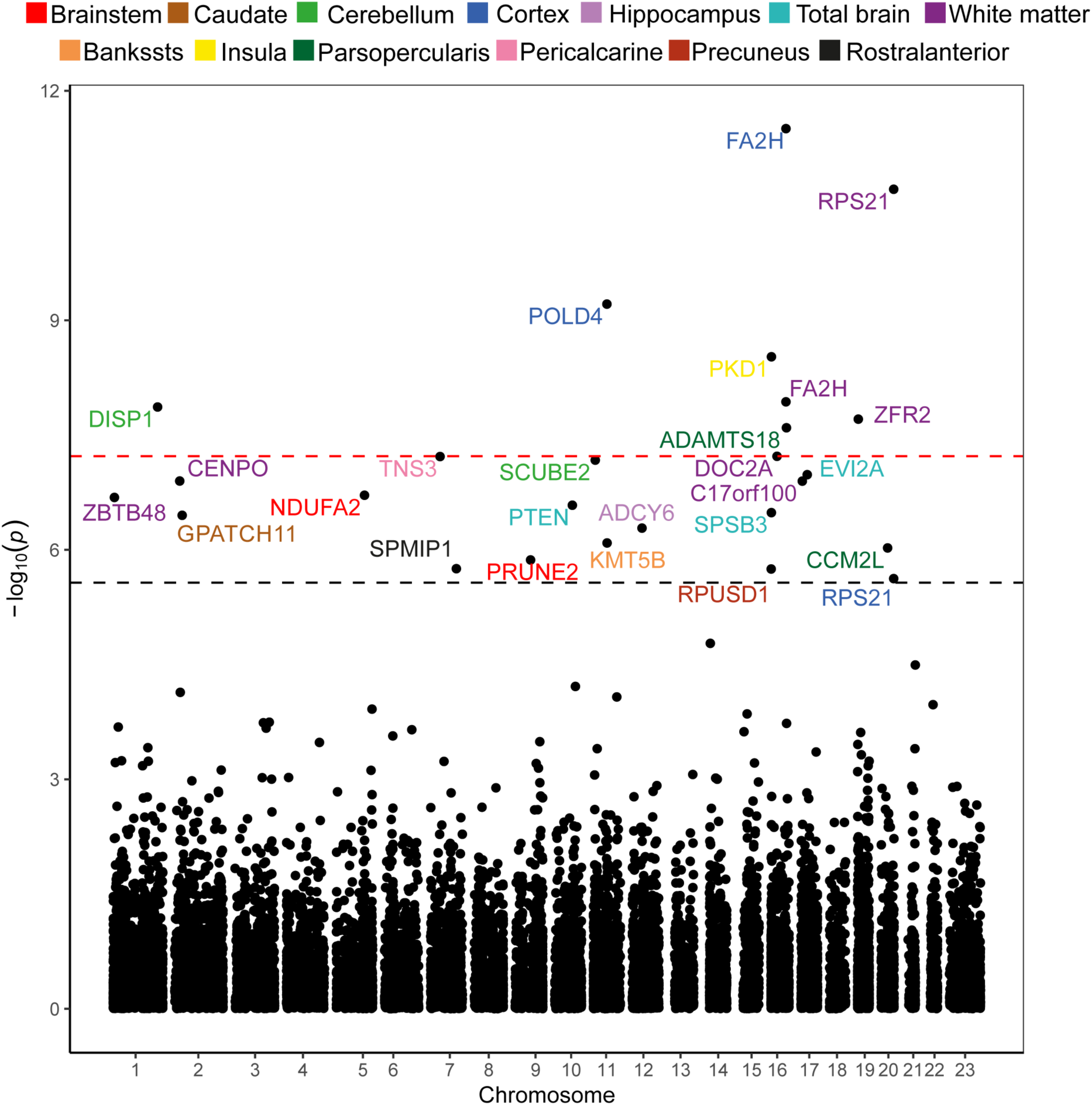
The pLOF+missense gene associations highlight 26 gene phenotype associations. The gene names are colour coded based on the phenotypes they were associated with. The red dashed line represents study wide significance (P<0.05/(18613*44)) and the black dashed line represents genome wide significance (P<0.05/18613). Bankssts represents Banks of the Superior Temporal Sulcus, Parsopercularis represents Pars opercularis, Rostralanterior represents rostral anterior cingulate cortex, Cortex represents cortical grey matter. Gene associations are available in **Supplementary Table 2**.

Seven of the 24 unique genes in the pLOF+missense analyses have been linked to brain development phenotypes in ClinVar (**Table 1**; **Supplementary Table 2**). None of the SWS/GWS genes in the synonymous analyses were linked to brain related phenotypes. This suggests that compared to the negative control, the genes associated with brain volume through pLOF and missense variants appear to be relevant for healthy brain formation and development. Full discussion of the pLOF+missense associated genes and their evidence in previous literature is available in the **Supplementary Note**. While seven genes were linked with rare brain disorders, four of these seven (*DISP1*, *SCUBE2*, *PTEN*, *FA2H*) had the most evidence within this study and in previous literature.

## *DISP1*, *SCUBE2*, and cerebellar volume

Two genes, *DISP1* and *SCUBE2*, were associated with smaller cerebellar volume through pLOF+missense variants. This is noteworthy because *SCUBE2* and *DISP1* work together in the same pathway to regulate secretion of sonic hedgehog protein^14,15^, a key signalling molecule for the development of the nervous system. While we could not find previous literature to support the role of *DISP1* and *SCUBE2* in cerebellar volume specifically, they have been connected to rare brain disorders. Mutations in *DISP1* and *SCUBE2* have been previously associated with holoprosencephaly^14,16^, a cerebral malformation with high phenotypic variability. Interestingly, holoprosencephaly has shown a complex inheritance where variants in multiple genes (*DISP1*/*SCUBE2*/*SHH*) or compound heterozygous variants in the same gene are needed in order to produce the disease phenotype^16,17^.

The combined mutations required to cause holoprosencephaly are unlikely to be observed in our sample (UKB) because it does not contain individuals diagnosed with holoprosencephaly. We searched within our sample to determine if any holoprosencephaly associated mutations in *SCUBE2*, *DISP1*, or *SHH* co-occured in the same individuals. We found that only one pair of variants (p.Met1096Thr in *DISP1* and p.Gly222Ser in *SCUBE2*) were present in the same individuals (**Supplementary Table 3**; **Supplementary Note**). This combination of variants only co-occurred in four individuals. One of these variants (p.Met1096Thr) has been shown in a previous study^16^ to be insufficient to cause holoprosencephaly alone but can cause holoprosencephaly when in combination with a mutation in the gene encoding sonic hedgehog (*SHH*). The four double carriers showed a small (non-significant) decrease in cerebellar volume but not in total brain volume compared to non-carriers or single carriers. This suggests that these variants may be additive to decrease cerebellar volume but not to a pathological degree. Evidence in this study and previous work suggests that single variants in *SCUBE2* and *DISP1* are insufficient to cause severe disorders, with compound heterozygous mutations or mutations in *SHH* required for severe phenotypes. We show that single variants in *SCUBE2* and *DISP1* are sufficient to cause more moderate phenotypes like lower cerebellar volume. This adds support for the importance of the *SHH* pathway in neurodevelopment and brain health.

## *PTEN* and macrocephaly

We found that a burden of pLOF and missense variants in *PTEN* were associated with larger total brain volume. *PTEN* is known to influence cell growth and survival through the PI3K and mTOR pathways^18^ and has been linked to multiple disease including cancer^19^ and macrocephaly^20,21^. The association with total brain volume in our study was largely driven by three singletons (variants only present in one individual), which explained between 35% and 20% of the gene association alone. All three of these variants were present in different individuals. The variant which explained most of the association signal was a stop gained variant (p.Arg335Ter in ENST00000371953) that was characterised as pathogenic for various cancers and macrocephaly in ClinVar. This variant was previously observed in a 7 year old male with an occipital–frontal circumference more than 3.5 standard deviations larger than the mean, along with developmental delay and benign neoplasms^20^. Additionally, one (p.Tyr68His in ENST00000371953) of the other two variants was characterised as pathogenic for various cancers and macrocephaly in ClinVar and a previous study reported that a four year old child with the p.Tyr68His mutation was observed with an occipital–frontal circumference 5.3 standard deviations larger than the mean^21^. In our sample, the individuals carrying the p.Arg335Ter and p.Tyr68His variant had a total brain volume 2.53 and 2.22 standard deviations larger than the median total brain volume respectively. This suggests that *PTEN* mutations are reliably linked to macrocephaly in children and adults across clinical and population-based cohorts.

## *FA2H* and white matter

One significant gene (*FA2H*) showed an opposing association with white matter and cortical grey mater volume; where a burden of pLOF and missense variants were associated with lower white matter volume and associated with higher cortical grey matter volume. The same variant (p.Glu78Lys in ENST00000219368) was prioritised for both phenotypes and this variant also showed the opposing effect direction. Based on previous literature, *FA2H* has more evidence for being linked to white matter volume than cortical grey matter volume. *FA2H* encodes fatty acid 2-hydroxylase, an enzyme that produces fatty acids for myelin production^22^. Mutations in *FA2H* (c.786+1G>A and p.Asp35Tyr) have been associated with a progressive neurologic disorder which presents with spasticity, dystonia, and white matter degeneration^22^. While previous literature supports the association of *FA2H* with white matter volume, we were unable to find literature connecting *FA2H* to cortical grey matter volume. It is likely that the positive association with the cortical grey matter was caused by conditioning on total brain volume; causing an opposing effect direction. When total brain volume was removed as a covariate the association of *FA2H* with Cortical grey matter almost disappeared (P=3.12×10^-12^ to P=0.045) whereas the association with white matter did not (P=1.16×10^-8^ to P=3.85×10^-4^) (**Supplementary Note**). These results and prior research suggest that the association with cortical grey matter is a statistical artefact of the influence on white matter. This study and previous studies suggest that mutations in *FA2H* are linked to lower white matter volume through affecting myelin maintenance.

## Convergence with common variants

In addition to highlighting genes associated with variation in brain volume and rare brain disorders, we were interested in whether the rare variant associations with brain volume differences would converge with common variant associations with brain volume differences (**Supplementary Note; Supplementary Table 4**). We used two strategies to assess the convergence between rare and common variants. First, we assessed the gene significance in an independent dataset that studied brain volume using common variants (ENIGMA2^5–7^). We chose the ENIGMA2 results over more recent ENIGMA studies as recent ENIGMA studies included UKB participants so these datasets would not be independent from this study. Second, since only 7 phenotypes overlapped across the ENIGMA2 study and our study, we also looked for convergence between common and rare variants using the same set of people in the UKB.

In the ENIGMA2 results, no pLOF+missense or synonymous genes were GWS (P<0.05/18613) and the genes from the pLOF+missense analyses did not nominally replicate (P<0.05) at a higher rate than the genes from the synonymous analyses (pLOF+missense: 9/24; synonymous: 3/10). This could be due to the low overlap in phenotypes so only 5 pLOF+missense genes could be assessed in the equivalent phenotype in the ENIGMA2 data. As expected there was more similarity in the rare and common variant results within the UKB than between the UKB and ENIGMA2 data (**Supplementary Note; Supplementary Table 4**). While none of the 24 pLOF+missense genes were GWS (P<0.05/18613) in the ENIGMA2 results, there were 4 GWS genes in the UKB common variant results. However, like with the ENIGMA2 results, the pLOF+missense genes did not replicate (P<0.05/18613) at a higher rate than the synonymous genes (pLOF+missense: 4/24; synonymous: 2/10).

Notably, one gene (*SCUBE2*) showed an association with lower cerebellar volume in the common and rare variant analyses, potentially through independent mechanisms (**Figure 3**). We found that a rare missense variant in exon 6 (p.G222S; MAF=9.53×10^-4^) and a common variant downstream of exon 17 (rs11042158; MAF=0.40) were linked with lower cerebellar volume in the rare and common analyses respectively. These associations likely represent two separate associations as these variants were in extremely low linkage disequilibrium (R^2^=0.0013) (**Supplementary Note; Supplementary Table 5)**. As expected, the common variant showed a smaller effect on cerebellar volume compared to the rare missense variant (rs11042158 beta= -517.30; pG222S beta= -3480.55; the beta represents mm^3^ after conditioning on total brain volume and other covariates). The C allele of the common variant was linked with lower cerebellar volume in our study and lower *SCUBE2* expression in the cerebellar GTExv8 data^23^, which suggests that this variant may impact cerebellar volume through impacting *SCUBE2* expression. These results suggest that the common and rare variant associations in *SCUBE2* converged to cause a decrease in cerebellar volume through independent mechanisms (decreased expression and change of the amino acid sequence).

**Figure 3:**
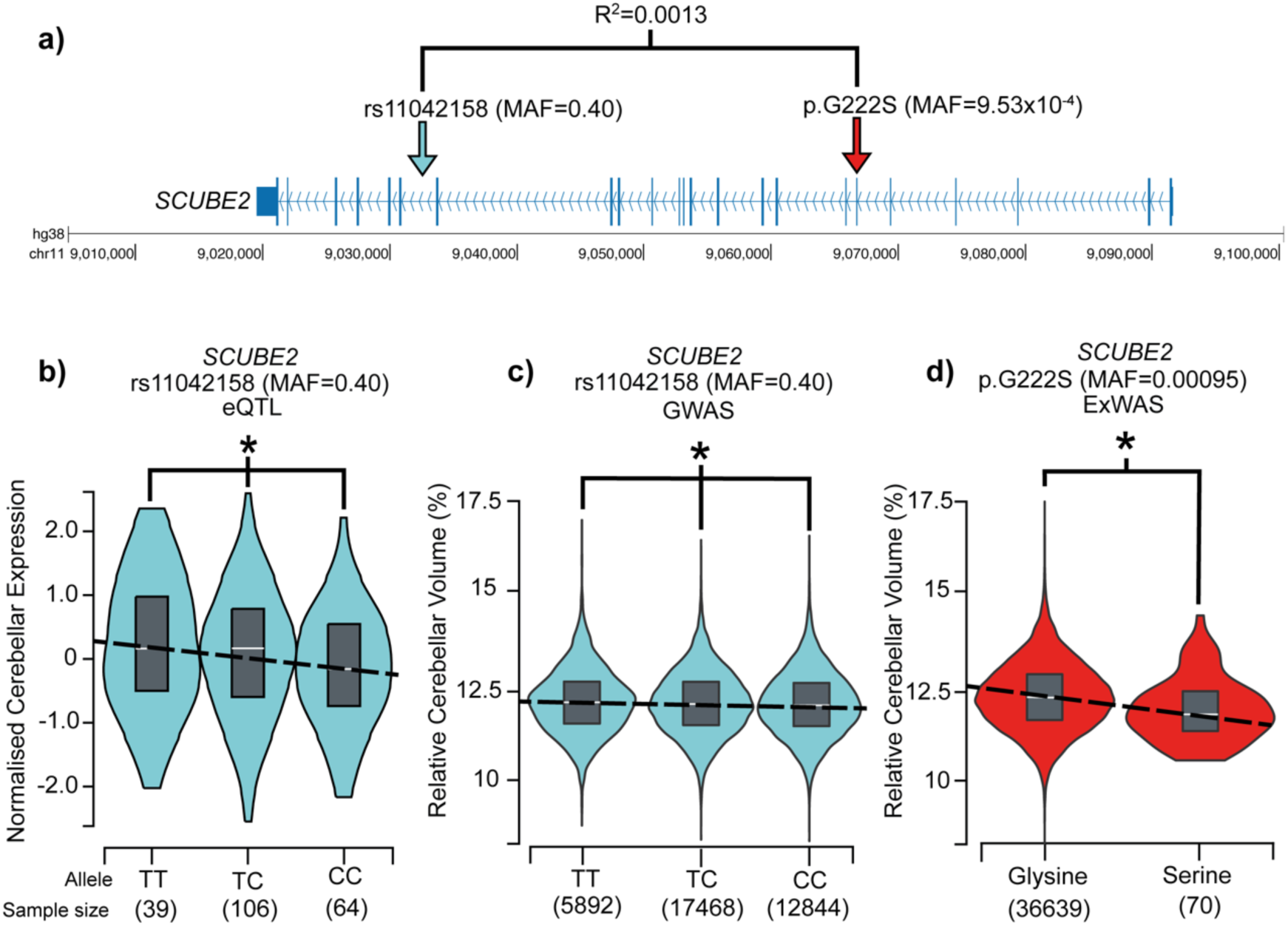
a) A common variant (rs11042158) and a rare variant (p.G222S) in SCUBE2 were independently associated with cerebellar volume in the same set of individuals. R^2^ represents the squared correlation between the variants. MAF represents the minor allele frequency. b) The C allele of rs11042158 is significantly associated with decreased expression of SCUBE2 in the GTEx v8 cerebellum^23^. The asterisk represents that the slope of the dashed line was significantly different from 0 (P=0.00011). The white line within the box plot represents the median normalized expression for each genotype. c) The C allele of rs11042158 is significantly associated with lower cerebellum volume in the UK Biobank sample. The asterisk represents that the slope of the dashed line was significantly different from 0 (P=6.94×10^-13^). The Y axis is the relative cerebellar volume which represents the percentage of the total brain volume that is made up by the cerebellum. The white line within the box plot represents the median. d) Serine at amino acid position 222 of SCUBE2 (ENST00000649792) is significantly associated with lower cerebellum volume in the UK Biobank sample. The asterisk represents that the slope of the dashed line was significantly different from 0 (P=2.20×10^-3^). The Y axis is the relative cerebellar volume which represents the percentage of the total brain volume that is made up by the cerebellum. The white line within the box plot represents the median.

## Discussion

In this study we performed exome wide association analyses for 44 different brain volume phenotypes in the UKB. While utilising rare variants in this cohort had limitations (**Supplementary Note**), we were able to highlight 7 genes associated with brain volume differences which are known to be linked to brain disorders through rare variants. Three of these genes were involved in fundamental pathways related to body plan development (*SCUBE2*/*DISP1* in sonic hedgehog pathway^14,15^) and cell metabolism/growth (*PTEN* in mTOR pathway^18^). Interesting, Klein et al. (2019)^24^ suggested that the sonic hedgehog pathway and mTOR pathways interact, which explains the similar brain volume phenotypes in patients with mutations in other genes in these pathways. This suggests that other genes in these pathways may be identified in future rare variant association analyses. While deleterious mutations in these pathways can lead to severe brain disorders (holoprosencephaly), we show that missense and pLOF variants in these same genes can lead to more moderate phenotypes, like sub-clinical differences in brain volume. This suggests that the specific mutation or the combination of mutations (in the case of holoprosencephaly) are important in determining whether an individual develops a severe or moderate phenotype.

In addition to highlighting genes associated with brain disorders and variation in brain volume, we also showed that common and rare variants can converge on the same gene (*SCUBE2*) to influence a phenotype (cerebellar volume) through independent mechanisms (decreased expression and amino acid substitution). We also showed that, expectedly, the common variant eQTL showed a considerably lower effect size than the rare missense variant. We also highlighted an example where mutations in two genes in the same pathway were linked to the same phenotype, where *SCUBE2* and *DISP1* were both associated with cerebellar volume and are known to modulate sonic hedgehog signalling. We replicated a finding in adults which linked mutations in *PTEN* to macrocephaly in children and provided support for the link between *FA2H* and white matter volume. Gene-set and tissue enrichment analysis also identified expected associations like the association of myelin maintenance with white matter volume and that genes enriched in association signal for total brain volume were overexpressed in the brain (**Supplementary Note**). Overall, we were able to use rare variant data to explore genetic contributors to differences in brain volume and contextualise these results using previous literature on rare brain disorders. These results help us to understand how genes associated with rare brain disorders can contribute to brain variability and can be used to explain phenotypes in unsolved cases where patients present with multiple genetic variants and multiple phenotypes.

## Methods

### Sample overview

We used exome sequencing data from 40,374 UKB participants of European ancestry who had MRI data available. The UKB is a large population-based biobank which includes 503,325 individuals. Individuals were selected for participation between 2006 and 2010. Invited individuals were between 40 and 69 years old, registered with the National Health Service, and living within 25 miles of one of the study research centres. Various data were collected from the individuals, including questionnaire answers, medical records, body measurement, and genetic data. A total of 40,374 (53.6% female) participants of European ancestry were included in this analysis. The median age was 65 at the time of imaging.

Approximately 15% of the included participants had a third degree relative included in the analysis. All participants provided written informed consent. The UKB received ethical approval from the National Research Ethics Service Committee North West-Haydock (reference 11/NW/0382). All study procedures were in accordance with the World Medical Association for medical research. Access to the UK Biobank data was obtained under application number 16406. The genotyping and exome sequencing protocols are described in Bycroft *et al.* (2018)^25^ and Backman *et al.* (2021)^11^. Array genotypes were on build GRCh37 and exome sequencing data was on build GRCh38.

### Phenotypes and covariates

We created 44 phenotypes which included total brain volume and 43 bilateral brain regions. Global and subcortical volumes were extracted from the aseg segmentation files^26^ and cortical volumes were parcellated according to the Desikan-Kiliany^27^ atlas in the Freesurfer processing performed by the UKB team^28^. The subcortical regions included in this study were the thalamus, caudate, putamen, pallidum, hippocampus, amygdala, nucleus accumbens, and brainstem. Bilateral volumes (all except brainstem, total cerebellar volume and total brain volume) were averaged between the left and the right hemisphere of each subject. In the context of this study, total brain volume includes the cerebellum and excludes vessels, dura and ventricles. Supplementary analyses of the ventricles and intracranial volume are available in the **Supplementary Note**. Cerebellar volume includes both grey and white matter in the cerebellum. White matter volume includes all white matter in the cerebrum. Cortical grey matter volume includes all the grey matter between the pial and white surfaces, except subcortical elements inside of the cortical ribbon. With respect to regional volumes, cortical and subcortical volumes refer to the volume of grey matter segmented in each of the respective regions.

We used the same set of covariates for all of the analysis, except the total brain analysis which did not include total non-ventricular brain volume as a covariate. The covariates used were sex, age at imaging, age at imaging squared, handedness, total non-ventricular brain volume, population principal components (PCs) 1-20, scanning site, table coordinates, coil position, amount of linear and non-linear warping in T1 registration, discrepancy between T1 and registration template after registration, and exome batch. As sensitivity analyses, we analysed the significant genes from the regional volumes without including total brain volume as a covariate (**Supplementary Note**). We used array-based genotypes to empirically assign individuals to ancestral continental populations using the 1000 Genomes reference panel based on Mahalanobis distance of the first 20 PCs^29^. PCs were created using FlashPCA2^30^.

We performed quality control by removing individuals who had a value greater than five times the median absolute deviation from the overall median of the non-linear warping in T1 registration and discrepancy between T1 and template after registration^31^. We also removed outlier individuals who had a phenotype value more than 4 standard deviations from the mean (in both directions) to avoid extreme phenotypes biasing the results (**Supplementary Note**). If an individual was an outlier in any phenotype, they were removed from all analyses. This removed 727 individuals. After removing these individuals, along with individuals who chose to remove themselves from the UKB cohort, we had a set of 40,374 individuals with total brain volume measures and exome sequencing data, and 36,709 individuals with regional brain volume measures and exome sequencing data.

### Variant annotation

Initially, we annotated the entire set of variants available in the UKB exome sequencing data. Before annotation we removed variants where less than 90% of all genotypes for that variant had a read depth less than 10 (ukb23158_500k_OQFE.90pct10dp_qc_variants.txt). Variants were annotated with Ensembl variant effect predictor (VEP) v100.432^32^ using Ensembl version 100 data. The variants were annotated with respect to the minor allele (least frequent). We restricted all of our analyses, except the synonymous analyses, to only missense and predicted LOF (pLOF) variants based on these annotations. The variants were included if they were missense or pLOF in any of the transcripts included in Ensembl v100. pLOF variants were defined as having the following consequences: start_lost, stop_lost, frameshift_variant, stop_gained, splice_donor_variant, splice_acceptor_variant, and transcript_ablation. To create a high confidence and low confidence set of variants, we annotated pLOF variants with the LOFTEE plugin (github commit 2df8880)^33^ and we annotated missense variants with the REVEL v1.3 plugin^34^. Any missense variants with a REVEL score >50 were considered high confidence, any variants with a REVEL score between 0 and 50 were considered low confidence, any pLOF variants defined as high confidence by LOFTEE were considered high confidence and all other pLOF were considered low confidence. With these annotations we created two masks; “high confidence” which included the high confidence missense and pLOF variants and “all variants” which included all pLOF and missense variants.

### Gene based analyses

We performed gene-based analyses using REGENIE v3.4.1^35^ using the two step procedure. First, we created genomic predictions (step 1) using typed variants extracted from the array-based genotypes. Only autosomal typed variants with MAF>0.01, MAC>100, genotyping rate >0.9, and Hardy-Weinberg equilibrium P-values >1×10^-15^ were included in step 1. We created these predictions separately for the total and regional brain volume analyses resulting in two sets of predictions. Then in step 2, we used those predictions, along with the exome sequencing data in BGEN format, and annotation files to perform gene-based analyses. We only included variants that were included in the two masks (high confidence pLOF+missense and all pLOF+missense) and variants with a MAF<0.01 in the subset of individuals being analysed (40,374 individuals in the total brain volume analysis and 36,709 individuals in the regional brain volume analyses).

We analysed the genes using multiple methods which were then combined in the GENE_P approach implemented in REGENIE (--rgc-gene-p). This method performs burden, kernel, and ACAT tests (--vc-tests skato,acato-full) and then combines the resulting P-values using aggregated Cauchy association tests. This results in a single P-value for each gene after aggregating evidence across multiple test types, allele frequencies, and variant masks (high confidence and all variants). In total, 24 association tests were performed for each gene and then combined to create a single GENE_P value. These tests were:

1. Burden of high confidence missense and pLOF singletons
2. Burden of high confidence missense and pLOF variants with MAF<0.0001
3. Burden of high confidence missense and pLOF variants with MAF<0.001
4. Burden of high confidence missense and pLOF variants with MAF<0.01
5. Burden of all missense and pLOF singletons
6. Burden of all missense and pLOF variants with MAF<0.0001
7. Burden of all missense and pLOF variants with MAF<0.001
8. Burden of all missense and pLOF variants with MAF<0.01
9. ACAT combination of burden tests
10. SBAT combination of burden tests
11. SBAT combination of burden tests of variants with positive effects
12. SBAT combination of burden tests of variants with negative effects
13. ACAT combination of high confidence missense and pLOF variants (ACATV)
14. ACAT combination of all missense and pLOF variants (ACATV)
15. SKAT analysis of high confidence missense and pLOF variants
16. SKAT analysis of all missense and pLOF variants
17. SKAT-O analysis of high confidence missense and pLOF variants
18. SKAT-O analysis of all missense and pLOF variants
19. ACAT combination of the SKAT-O, burden, and ACATV analyses using high confidence missense and pLOF variants
20. ACAT combination of the SKAT-O, burden, and ACATV analyses using all missense and pLOF variants
21. ACAT combination of the burden and ACATV analyses using high confidence missense and pLOF variants
22. ACAT combination of the burden and ACATV analyses using all missense and pLOF variants
23. ACAT combination of the burden and ACATV analyses
24. ACAT combination of the burden and SKAT-O analyses

Additionally, we specified that all variants with a MAC<5 should be aggregated before inclusion in the kernel tests. We repeated this analysis process using only synonymous variants as a negative control. Synonymous variants were any variant annotated with “synonymous_variant” by VEP.

We defined genome wide significant (GWS) genes as genes with a GENE_P P-value less than 0.05 after Bonferroni correction for the number of genes tested in that phenotype (0.05/18613= 2.69×10^-6^). We also defined study wide significant (SWS) genes as genes with a GENE_P P-value less than 0.05 after Bonferroni correction for the number of genes tested in all phenotypes (0.05/(18613*44)= 6.11×10^-8^). Genomic inflation factors were calculated on the GENE_P using the qchisq() function in R v4.2.1^36^.

### Gene-set analyses

We performed gene-set analyses using MAGMA v1.10^37^ and the output of the gene-based analyses. We formatted the REGENIE output into the magma raw file format, where we assumed there was no linkage disequilibrium (LD) between variants. This required calculating the number of variants mapped to each gene and estimating the Z-score from the P-value using the qnorm() function in R v4.2.1^36^. Then we performed gene-set analyses using the MSigDB v2023.1Hs gene-sets (c2 and C5.GO). SWS and GWS gene-sets were defined as P<0.05/(17018*44) and P<0.05/(17018), where 17,018 gene-sets were tested. Tissue enrichment analyses were performed using the same MAGMA raw files and GTEx^23^ v8 average gene expression. The model was one-sided (--model direction=greater) and the association was conditioned on the average gene-expression of all tissues (condition-hide=Average). SWS and GWS associations were P<0.05/(54*44) and P<0.05/54, where 54 tissues were tested. Significant brain regions associations were conditioned on the expression of general brain expression which was the average expression of GTEx v8 brain tissues.

### Comparison to common variant results

ENIGMA2 common variant GWAS summary statistics for intracranial volume (ICV)^7^, accumbens^5^, amygdala, caudate, hippocampus^6^, putamen, and thalamus were download and MAGMA gene analyses were performed using MAGMA v1.10^37^. Gene boundary definitions (GRCh37) were obtained from Biomart^38^ using Ensembl gene IDs. The 1000 genomes^29^ Europeans LD information was used during this analysis and this was obtained from the MAGMA website (https://cncr.nl/research/magma/). Common variant GWAS in the UKB were performed for total brain volume and all 44 regional brain volume phenotypes using the same individuals as the rare variant analyses. The analysis was performed like the rare variant analyses except they only included variants with MAF >0.01, exome batch was replaced by array batch as a covariate, and variants were analysed alone (not aggregated within genes). The imputed genotypes in plink bed format were used for the regression, with variants with INFO <0.9 being excluded. The resulting GWAS summary statistics were used for MAGMA gene analysis using the same method as the ENIGMA MAGMA analysis above. LD between rare and common variants were estimated on the UKB individuals using PLINK v1.90b6.26^39^ (--r2).

### Variant prioritisation

Variants within GWS and SWS genes were prioritised by performing leave-one-variant-out (LOVO) analyses. LOVO analyses were performed using the test types where leaving variants out is possible (burden, SKAT, and ACATV tests). The significance of the gene after removal of a variant was compared to the significance of the most significant burden, SKAT, or ACATV test for that gene. Variants that explained more than 10% of the gene significance were prioritised for further investigation. Additionally, we performed single variant association in REGENIE v3.4.1 of all missense and pLOF variants in GWS and SWS genes to investigate the effect directions and significance of variants in those genes.

## Data availability

All gene level pLOF+Missense summary statistics for all phenotypes are available for download at https://github.com/dwightman/UKB_Exome_BrainVolume. Genotype level data from the UK Biobank can be accessed through application https://www.ukbiobank.ac.uk/enable-your-research/apply-for-access. ENIGMA2 data can be obtained from https://enigma.ini.usc.edu/research/download-enigma-gwas-results/. GTEx data for tissue enrichment analyses can be obtained from GTEx https://gtexportal.org/home/downloads/adult-gtex/overview. Gene-set definitions can be obtained from MSigDB https://www.gsea-msigdb.org/gsea/downloads.jsp.

## Code availability

Analysis code is available at https://github.com/dwightman/UKB_Exome_BrainVolume.

## Acknowledgements

DP was funded by NWO Gravitation: BRAINSCAPES: A Roadmap from Neurogenetics to Neurobiology (Grant No. 024.004.012), and a European Research Council advanced grant (Grant No, ERC-2018-AdG GWAS2FUNC 834057).

DW was funded by NWO Gravitation: BRAINSCAPES: A Roadmap from Neurogenetics to Neurobiology (Grant No. 024.004.012).

BAPCM was funded by NWO Gravitation: BRAINSCAPES: A Roadmap from Neurogenetics to Neurobiology (Grant No. 024.004.012).

MPvdH was supported by NWO VIDI Grant 452-16-015 and the ERC Consolidator of the European Research Council Grant 101001062.

## Author information

**Department of Complex Trait Genetics, Center for Neurogenomics and Cognitive Research, Amsterdam Neuroscience, Vrije Universiteit Amsterdam, Amsterdam, The Netherlands**

Douglas P. Wightman, Bernardo A.P.C. Maciel, Rachel M. Brouwer, Martijn P. van den Heuvel, Danielle Posthuma

**Department of Child and Youth Psychiatry, Emma Children’s Hospital, Section Complex Trait Genetics, Amsterdam Neuroscience, Amsterdam UMC, 1081 HV Amsterdam, The Netherlands.**

Martijn P. van den Heuvel, Danielle Posthuma

D.P.W. performed the analysis, interpreted the results, and wrote the manuscript.

B.A.P.C.M. created the phenotypes and interpreted the results. M.P.v.d.H, R.M.B, and D.P. provided supervision and feedback.

No authors have any competing interests.

