## Supplementary Note for "Rare variant aggregation highlights rare disease genes associated with brain volume variation"

### Supplementary Methods

#### Intracranial and ventricle volume analyses

First ventricle and intracranial volume (ICV) measures were extracted from the UKB data using the following data fields: 26554, 26555, 26585, 26586, 26521. Any missing data in instance 2 of these fields was replaced with the value from instance 3. The left ventricle volume was calculated as the combination of the “Volume of Lateral-Ventricle” and “Volume of Inf-Lat-Vent” (26554+26555). The right ventricle was calculated in the same way using these data fields: 26585+26586. Then the ventricle volume was calculated as the mean of the left and right ventricles. Individuals with a phenotype value 4 standard deviations from the mean were removed and the individuals were restricted to the same set of 36,709 individuals used in the regional brain volume analyses. The same two step procedure, using REGENIE v3.4.1[1], described in the main text was used to analyse the ventricle volume. Genomic predictions (step 1) were created using typed variants extracted from the array-based genotypes. Only autosomal typed variants with MAF>0.01, MAC>100, genotyping rate >0.9, and Hardy-Weinberg equilibrium P-values $>1x10^{-15}$ were included in step 1. Then in step 2, we used those predictions, along with the exome sequencing data in BGEN format, and annotation files to perform gene-based analyses. We only included variants that were included in the two masks (high confidence pLOF+missense and all pLOF+missense) and variants with a MAF<0.01 in the subset of individuals with ventricle volume. We used the same covariates as the main text except ICV was used instead of total brain volume.

As a sensitivity analysis we examined the significance of *PTEN*, *EVI2A*, and *SPSB3* with ICV by using the same analysis plan as total brain volume. Intracranial volume was taken from data field 26521 and people with phenotype values 4 standard deviations from the mean were removed. This left 37,458 individuals. Then the gene analysis was performed in the same way as described in the main text for the total brain volume analysis.

#### Prioritised variant comparison to ClinVar

We compared the CADD score of variants prioritised in our LOVO analyses to variants in ClinVar in the same genes. We only compared variants in the 7 genes which had ClinVar associations with a brain related phenotype (*FA2H*, *DISP1*, *SCUBE2*, *NDUFA2*, *PTEN*, *GPATCH11*, *KMT5B*). We selected pathogenic variants in those genes associated with a brain related phenotype in ClinVar to compare with the variants prioritised in LOVO analyses (**Supplementary Table 15**). We annotated the variants with CADD scores (a measure of deleteriousness) using the VEP[2] web interface (GRCh38), where we queried the variants in VCF format. The median and mean CADD score for each gene was calculated across the variants in that gene. *KMT5B* was not included in the comparison because the variant linked to a brain related phenotype in *KMT5B* in ClinVar did not have an associated CADD score in VEP.

### Supplementary Results

#### Global inflation factors

The genomic inflation factors of the pLOF+missense analyses ($\lambda$) were generally low (less than 1), with a range of 0.82 in the isthmus cingulate to 0.95 in the brainstem (**Supplementary Table 1**). These low values were expected given the relatively low sample size to identify rare variants (~30-40k individuals) and the restriction of limiting the analysis to missense and pLOF variants. The genomic inflation factors of the synonymous variants were on average 1.16x larger than the pLOF+missense analyses (0.88-1.02), but the pattern was similar to the pLOF+missense analysis (Pearson correlation=0.76). This was expected given that pLOF and missense variants are more likely to be selected out of the population leaving lower spread of association signal (lower genomic inflation factor) compared to the neutral synonymous variants which should distribute signal according to the null model ($\lambda$=1).

#### *DISP1*, *SCUBE2*, and *SHH*

Mutations in *DISP1* and *SCUBE2* have been previously associated with holoprosencephaly[3,4], a cerebral malformation with high phenotypic variability. Holoprosencephaly has shown a complex inheritance where variants in multiple genes or compound heterozygous variants in the same gene are needed in order to produce the phenotype[3,5]. A previous study[3] showed that mutations in *DISP1* were insufficient to cause holoprosencephaly alone but caused holoprosencephaly when in combination with a mutation in *SHH*. We queried our sample to look for the co-occurance of mutations mentioned in previous literature or prioritised in this study in *DISP1*, *SCUBE2*, and *SHH* **(Supplementary Table 3**).

Expectedly, we did not find any carriers of the *SHH* mutations mentioned in Kim *et al.* (2019)[4]. We also did not observe any carriers with more than one mutation for the other variants discussed in Mouden *et al.* (2016)[3] or Kim *et al.* (2019)[4] in *SCUBE2* or *DISP1*. We did observe 4 individuals who were carriers of p.Met1096Thr in *DISP1* and p.Gly222Ser in *SCUBE2*. Both of these variants were the prioritised variants for these genes in this study; p.Met1096Thr was also reported in Mouden *et al.* (2016)[3]. The median cerebellar volume for these individuals was smaller than the median cerebellar volume for non-carriers (-0.22 stdev), p.Met1096Thr only carriers (-0.0025 stdev), and p.Gly222Ser only carriers (-0.070 stdev). The difference between the cerebellar volumes of the double carriers was not significantly different from the cerebellar volumes of the other carriers and non-carriers, due to the low sample size of the double carriers (N=4). However, the pattern observed with cerebellar volume was not observed with total brain volume, where carriers of both variants had larger brains overall compared to non-carriers (+0.020 stdev) and *DISP1* carriers (+0.065 stdev), but smaller brains than *SCUBE2* carriers (-0.15 stdev). Additionally, *SCUBE2* and *DISP1* were not associated with any other phenotype ($<1x10^{-4}$) (**Supplementary Table 6**) which suggests the association was specific to the cerebellum. Overall, individuals within our relatively healthy cohort (who do not have holoprosencephaly) were carriers of variants in *SCUBE2* and *DISP1* that cause brain volume changes but were not carriers of variants in both *SCUBE2* and *SHH* or *DISP1* and *SHH*. This suggests that alone these variants cause moderate phenotypes and that in combination they can cause more severe phenotypes.

#### Convergence with common variant datasets

In the absence of a direct replication set, we looked for convergence of results in an independent dataset that studied brain volume using common variants (ENIGMA2). We found that 9 of the 24 pLOF+missense genes had nominal significance (P<0.05) in the ENIGMA2 common variant results and that 3 of the 10 synonymous genes had nominal significance (**Supplementary Table 4**). The similar rate of replication in the pLOF+missense and synonymous associated genes suggests that the replication was due to chance. This convergence test was not ideal because the ENIGMA2 data only studied intracranial, accumbens, amygdala, caudate, hippocampus, putamen, and thalamus volume. Of the pLOF+missense associated genes, only 5 genes were associated with a phenotype that was present (or analogous) in the ENIGMA2 data (total brain/intracranial: *EVI2A*, *PTEN*, *SPSB3*; caudate: *GPATCH11*; hippocampus: *ADCY6*). However, none of those were nominally significant in the ENIGMA2 data in the same phenotype. It is possible that the low replication rate could be due common and rare variants not always converging to the same genes; for example, mutations in *PTEN* that cause macrocephaly are rare so would not be picked up in the common variant analysis.

We also looked for convergence within the same set of individuals across rare and common variants (UKB common). There, as expected, we found a greater degree of similarity in results (**Supplementary Table 4**). We found that 3 pLOF+missense ExWAS genes were SWS (P<0.05/(18613*44)) and 1 pLOF+missense gene was GWS (P<0.05/18613) in the common variant results (*ADAMTS18*, *DOC2A*, *SCUBE2*, *EVI2A*). In the synonymous results, two genes (*PSMA3* and *PRKAR2A*) were significant in the rare and common analyses. The convergence rate across common and rare variants was roughly the same between the pLOF+missense and synonymous analyses (Rare=4/24; Common=2/10). In the pLOF+missense analyses, two genes were significant in the same phenotypes in the common and rare analyses (*DOC2A* in white matter and *SCUBE2* in cerebellum); however, no genes that were significant in the synonymous analyses were also significant in the common variant analyses in the same phenotype.

We were interested in whether the genes that converged across common and rare variants in the pLOF+missense analyses converged through independent mechanisms. *SCUBE2* was significantly associated with cerebellar volume in the pLOF+missense and common variant analyses. LOVO analysis prioritised a variant (GRCh38: 11:9066793:C:T) which explained ~20% of the *SCUBE2* association with cerebellum (**Supplementary Table 7**). This variant was a missense variant in exon 6 (p.Gly222Ser in ENST00000649792) and was also in very low LD with the most significantly associated common variants ($R^2$<0.002). The C allele of the most significant common variant (GRCh37: 11:9054161:C_T/rs11042158) was located downstream of exon 17. This variant was associated with a smaller cerebellum (beta=-516.30, P=6.94x$10^{-13}$) and the C allele was also associated with decreased expression of *SCUBE2* in cerebellum in GTEx v8[6] (beta=-0.21, P=0.005). The missense variant prioritised in the rare variant analyses was independent from the eQTL found in the common variant analysis ($R^2$=0.0013) (**Supplementary Table 5**). The common and rare variant results converge to suggest that decreased expression and alteration of the amino acid sequence of *SCUBE2* is linked to decreased cerebellar volume.

*DOC2A* was significantly associated with white matter volume in the pLOF+missense and common variant analyses. Using the leave-one-variant-out (LOVO), we prioritised a single variant (GRCh38: 16:30006906:C:T) which explained ~96% of the pLOF+missense *DOC2A* association with white matter (**Supplementary Table 7**). This variant appears to be independent from all of the variants included in the common variant analyses ($R^2$<2x$10^{-4}$) which suggests that the association in the rare and common variants are through independent variants (**Supplementary Table 5**). The common variant association signal was spread throughout the gene, with the most significant variants being towards the end of the gene. We were unable to find any mechanism to explain the common variant associations (splice site alteration, eQTL, missense variant).

#### Study limitations

While we find relatively robust associations and show that the analyses were enriched for brain related findings compared to our null model (synonymous variants), there were multiple considerations that needed to be addressed for a study of rare variants and brain imaging phenotypes. First, rare variant studies require large sample sizes to find reliable associations because rare variants inherently have fewer carriers of the minor allele. Due to the cost of exome sequencing and brain imaging, there was not an available direct replication sample. We chose to use previous literature and convergence with common variants to assess the likelihood of associations as a substitute for direct replication. However, this method prevents the validation of novel associations. Next, the low spread of association in pLOF and missense hinders the power of gene-set enrichment analysis. Another consideration is that individuals with extreme phenotype values may harbour extremely rare variants with large impacts on brain volume but including these individuals will increase the false discovery rate (exemplified in the **Supplementary Note: Outlier removal**). In our study, we chose to remove individuals with extreme phenotype values (>4 standard deviations from the mean) to prevent the high number of false positives because we lacked a direct replication dataset. This means that we likely missed relevant associations. Previous studies[7,8], chose to limit analyses to variants with higher expected allele counts to limit the false positive rate; however we chose to focus on all variants due to the low number of individuals with imaging data (N~40k). A consideration for all regional brain volume analysis is how to disentangle general and specific brain volume, as creating ratio traits can lead to false positives[9]. In this study we chose to condition on total brain volume to find regional specific effects but this did also introduce a likely false association between cortical grey matter and *FA2H*. We additionally considered the association of each gene without conditioning on total brain volume to show how total brain volume contributes to each association. Overall, despite the difficulties of studying rare variants associated with brain imaging traits, we were able show that our association results were enriched for finding brain relevant genes compared to our null model and that multiple gene associations were well supported by previous research.

#### Gene-set analysis

We performed gene-set analysis using the MSigDB v2023.1Hs (hallmark+C2+C5) defined gene-sets to see if association signals would aggregate within gene-sets to highlight relevant biological processes (**Supplementary Table 8**). We identified two SWS ($P<0.05/(17018*44)$) gene-sets associated with total white matter and pericalcarine cortex. The gene-set associated with total white matter volume was

“GOBP_CENTRAL_NERVOUS_SYSTEM_MYELIN_MAINTENANCE” and was driven by *FA2H* (P=1.16x10$^{-8}$) and *PTEN* (P=8.09x10$^{-4}$). The gene-set associated with pericalcarine cortex (HALMOS_CEBPA_TARGETS_DN) was a set of genes regulated by a posited tumour suppressor protein (C/EBP alpha) and was largely driven by the association of *INHBB* (P=8.13x10$^{-6}$). The connection between C/EBP alpha and pericalcarine volume is unclear.

In addition to the 2 SWS gene-sets, there were 5 GWS gene-sets associated with total brain, cortical grey matter, isthmus cingulate, medial orbitofrontal cortex, and post central gyrus volume. These five gene-sets were related to histamine, the previous myelin maintenance gene-set, oligodendrocyte differentiation, liver cancer, and fatty acid metabolism, respectively (**Supplementary Table 8**). Except the gene-set related to myelin maintenance, which was also driven by *FA2H* (P=3.12x10$^{-12}$) in the cortical grey matter, these 5 GWS gene-sets do not appear to be driven by a small number of genes, rather an aggregation of many genes. Of these results, the association of myelin maintenance with white matter volume fits well with our current understanding of the phenotypes, whereas the other 6 associations do not have support in previous literature.

We also performed the gene-set analysis using the synonymous association results as a null comparison. We found 6 GWS gene-sets using the synonymous variant association results. (**Supplementary Table 8**). These gene-sets were related to hypoxia in breast cancer, observational learning, catecholamine secretion, lewy bodies, pentose and glucuronate metabolism, O-glycan biosynthesis. All of these gene-sets appear to be driven by a large number of genes in aggregate, with no single gene in the gene-set having a P-value < 1x10$^{-5}$. There was more spread of association signal in the synonymous gene analyses (higher genomic inflation factors) than the pLOF+missense gene analyses so it was expected that more gene-sets would be driven by a larger number of genes in the synonymous analyses.

#### Tissue enrichment analysis

We performed tissue enrichment analysis to investigate if genes associated with the brain volume phenotypes are overexpressed in any of the GTEx v8 54 tissue types. We did not find any SWS associations (P<0.05/(54*44)) but we did find 5 GWS (P<0.05/54) associations (**Supplementary Table 9**). Cerebellum expression was associated with total brain volume ExWAS signal (P=4.44x10$^{-4}$), half of this association was lost after conditioning on general brain expression (P=0.023). This suggests that the genes associated with total brain volume are likely to be expressed in the brain generally rather than the cerebellum specifically. Brain anterior cingulate cortex, cortical grey matter, frontal cortex and hippocampus expression were associated with precentral gyrus volume ($P<7x10^{-4}$). Brain anterior cingulate cortex was completely colinear with general brain expression which suggests that this association was not specific to anterior cingulate cortex. About 40% of the hippocampus association ($P=7.81x10^{-4}$) was reduced after conditioning on total brain expression (P=0.011). About 25-30% of the associations of cortical grey matter ($P=6.89x10^{-4}$) and frontal cortex ($P=5.14x10^{-4}$) could be explained by total brain expression (P=0.0071 and 0.0037). This suggests some specific overlap between precentral gyrus volume and hippocampus and cortical grey matter expression. The lack of association of brain expression with the other phenotypes could be explained by the sparsity of association signal in the phenotypes (low genomic inflation factors). The tissue enrichment analyses using the synonymous variant ExWAS results showed one GWS association between fusiform volume and pituitary expression ($P=8.96x10^{-4}$), ~10% of which was lost after conditioning on general brain expression (P=0.0018). It appears that the pLOF+missense gene associations are overexpressed in various brain tissues but the specific brain region associations do not align with the brain volumes tested to find gene associations.

#### Comparison of variants identified in this study to ClinVar variants

We hypothesised that the variants prioritised based on association with brain volume would be less deleterious to the proteins than the pathogenic variants in those genes reported in ClinVar. We hypothesised this because ClinVar annotates variants related to human disease so we expected variants related to disease to be more deleterious than variants related to non-disease phenotypes when the same genes are considered. We annotated the variants prioritised in this study and the variants in ClinVar linked to brain disorders with a measure of deleteriousness (CADD Phred Score). With the exception of *DISP1* and *FA2H*, we found the median CADD score of the brain volume and brain disorder associated variants to be similar (**Supplementary Table 10**). This suggests that, across 4 out of the 6 genes, variants associated with disease were no more deleterious than the variants we found to be associated with differences in brain volume. In the remaining two genes (*DISP1* and *FA2H*), the CADD scores of the disease related variants were considerably higher that the brain volume associated variants. The CADD score of the variant in *DISP1* associated with holoprosencephaly in ClinVar was 44 and 23.6 for the variant associated with cerebellar volume. The CADD score for the variant in FA2H associated with cerebellar atrophy was 31 and 24.2 for the variant associated with white matter volume. While this is interesting, the number of variants associated with the disease in ClinVar were extremely low (<5) (except with *PTEN* with 53 variants) so it is difficult to draw conclusions from such a limited sample. Future higher-powered research may find further rare variant harbouring genes associated with differences in brain volume that will allow for more comprehensive comparison with disease related genes.

#### Pleiotropy across genes in pLOF+missense analysis

While there were 26 GWS gene phenotype associations, there were only 24 unique genes associated with at least one phenotype because *FA2H* was SWS in cortical grey matter and white matter volume (Cortical grey matter GENE_P=$3.12 \times 10^{-12}$; White matter GENE_P=$1.16 \times 10^{-8}$) and *RPS21* was SWS in white matter volume and GWS in cortical grey matter volume (Cortical grey matter GENE_P=$2.38 \times 10^{-6}$; White matter GENE_P=$1.94 \times 10^{-11}$). *FA2H* was associated with higher cortical grey matter volume and associated with lower white matter volume. The association with white matter was well supported in previous literature[10] but the association with cortical grey matter was not. After removing total brain volume as a covariate, the association with white matter remained (P=$3.85 \times 10^{-4}$) but the association with cortical grey matter did not (P=0.045). This suggests that the true association was with white matter and the association with cortical grey matter was due to reduced white matter. The cortical grey matter association can potentially be explained by a higher proportion of the total brain volume being taken up by cortical grey matter if white matter volume was lower, which would induce a positive effect direction after correction for total brain volume. The association of *RPS21* was driven by a singleton which was associated with higher cortical grey matter and white matter volume. Neither of the associations were nominally significant (P<0.05) when total brain volume was not used as a covariate. Additionally, there was no support for *RPS21* being associated with brain phenotypes in previous literature. This association was likely a false positive given that it was driven by one singleton and had no support in previous literature.

#### Outlier removal

Individuals with extreme phenotype values may cause spurious associations with variants unique to them. This may cause multiple variants to be falsely associated with a phenotype. In the main analyses, we removed individuals with phenotype values more than 4 standard deviations from the mean to avoid outliers biasing the results. To test whether this step was necessary, we repeated the synonymous variant analyses without removing phenotypic outliers to assess the impact of outliers on the false positive rate. A substantially higher false positive rate was observed when phenotypic outliers were not removed compared to the synonymous analyses after removing outliers (**Supplementary Table 11**).

This effect was especially strong in the thalamus, where the most extreme phenotypic outlier values were observed. Removing outliers in this region caused a drop from 16 GWS genes to 1 in the null model (synonymous variants). A similar effect was observed for the parahippocampal gyrus as well (14 GWS genes to 0). Removing these outliers likely caused relevant associations to be missed (false negatives), as individuals carrying rare variants with large effects can be filtered out. However, since we lacked a direct replication sample, we chose the more conservative approach to limit false positives.

#### Ventricular volume analysis

In our primary analyses we excluded sections of the brain filled with cerebrospinal fluid, which meant we did not include ventricular volume as a phenotype in this study. For completeness we analysed the ventricles using the same approach as the main analysis (**Supplementary Methods**), except we included intracranial volume as a covariate instead of total brain volume. We found that this analysis contained relatively low spread of association signal ($\lambda$=0.84) with 0 SWS associated genes but 5 GWS associated genes (**Supplementary Table 12**). None of these genes (*ARF6*, *TCEAL5*, *FLII*, *FBXW5*, and *SGPP1*) were linked to brain related phenotypes in ClinVar.

*ARF6* encodes a small G protein which is important for the regulation and morphology of neurons[11] and has been suggested to affect amyloid-beta production in Alzheimer's disease (AD)[12]. In our study, mutations in *ARF6* were associated with increased ventricular volume and AD is known to be associated with increased ventricular volume[13]. One singleton (GRCh38: 14:49894024:C:G) explained 52% of the gene association. The carrier of this variant had not been diagnosed with AD but had been diagnosed with schizophrenia. Schizophrenia has also been associated with enlarged ventricles[14]. Since this variant is a singleton, the carrier of this variant had been diagnosed with schizophrenia, and schizophrenia is associated with increased ventricle size, it is difficult to determine whether *ARF6* is casually related to increased ventricular volume. *SGPP1* is involved in the degradation of sphingosine-1-phosphate[15] and sphingosine-1-phosphate has been suggested to be relevant to a wide range of brain disorders including AD[16]. In our study, mutations in *SGPP1* were associated with increased ventricular volume. One singleton (GRCh38: 14:63727883:G:A) explained 13.67% of the gene association but the carrier of this variant did not have any mental disorder that could explain the association. It is possible that the carrier of this variant could be in the early stages of neurodegeneration but since this association was driven by one singleton and sphingosine-1-phosphate has been suggested to be relevant to such a wide range of brain disorders it is difficult to discern if the association between *SGPP1* and increased ventricular volume is causal. We were unable to find any previous literature connecting *TCEAL5* or *FLII* to ventricular volume or any brain related phenotype. Interestingly, they have both been reported as being involved in myogenic differentiation[17,18]; however, that does not appear to be relevant for ventricular volume. *FLII* has been associated with ventricular volume in a previous study using the same participants[19]. We were unable to find previous literature connecting *FBXW5* to brain phenotypes.

#### Intracranial volume analysis

In our analysis, we defined total brain volume as the sum of the regional volumes and did not include the ventricles. Often intracranial volume, which includes the ventricles, dura, brain, and cerebrospinal fluid around the brain, is used to assess genetic associations[20–22]. We performed an association analysis of intracranial volume to see if the findings for total brain volume were also associated with intracranial volume (**Supplementary Table 2**). The association with *PTEN* was unaffected (P_TBV=$2.61 \times 10^{-7}$; P_ICV=$2.72 \times 10^{-7}$), the association with *SPSB3* was slightly reduced (P_TBV=$3.27 \times 10^{-7}$; P_ICV=$2.15 \times 10^{-5}$), and the association with *EVI2A* was substantially reduced (P_TBV=$1.04 \times 10^{-7}$; P_ICV=0.0024). We were interested in whether the effect of the prioritised variant in *EVI2A* (17:31318487:T:C; p.Q176R in ENST00000462804) was similar across total brain volume and ICV, as this variant explained ~90% of the gene association with total brain volume. The effect sizes were fairly equivalent but the standard error was larger in the ICV phenotype (Total brain volume: Beta=-18780, SE=3300, P=$1.27 \times 10^{-8}$; ICV: Beta=-17542, SE=4840, P=$2.89 \times 10^{-4}$). This suggests that the effect of this variant is still present on ICV but appears to affect the cortical and subcortical regions more than the overall intracranial space.

#### Regional volume associations without total brain volume as a covariate

We included total brain volume as a covariate when analysing regional brain volumes to try to avoid finding genetic factors for total brain volume when looking at specific regions. To see how robust those associations were, we re-analysed the significant genes without total brain volume as a covariate (**Supplementary Table 2**). Nine of the 26 gene-phenotype associations were not nominally significant (P<0.05) when total brain volume was removed as a covariate; of those 9, 6 of the associations were with white matter and 2 were with cortical grey matter. Only one gene (*FA2H*) associated with cortical grey matter or white matter was nominally significant after removing total brain volume as a covariate and in cortical grey matter that gene was just nominally significant (P=0.045). This was likely due to the fact that cortical grey matter and white matter were the two largest regions of the brain with the median volume of each being around 40% of the median total brain volume. These results suggest that the associations in the smaller regions were more robust to the removal of total brain volume as a covariate.

#### Leave-one-variant-out analysis

We investigated the association of the GWS gene-phenotype pairs by performing leave-one-variant-out (LOVO) analyses. In order to do this, we identified which test type (burden, SKAT, ACATV), variant annotation group (high confidence or all variants), and allele frequency mask (<0.01,<0.001,<0.0001, singletons) had the lowest P-value in the burden, SKAT, and ACATV tests for each GWS gene-phenotype pair. Then we tested that model after leaving one variant out to assess how individual variants affected gene significance. We prioritised variants if their removal caused the overall gene significance to decrease by >10%. Using this metric, we prioritised 41 variants across 26 genes-phenotype pairs (**Supplementary Table 7**). *CCM2L* was the only tested gene where no variant explained more than 10% of the gene association. For the rest of the genes, there was 1-3 prioritised variants. There were 10 genes where a variant explained more than 90% of the gene association and 9 genes where no variants explained more than 50% of the gene association. Overall, the LOVO analysis managed to highlight variants which explain a large amount of gene association in most cases.

#### Comparison to other rare variant studies

Three previous studies have looked at rare variant exome analyses of MRI based phenotypes[7,8,19] and in all three they studied a wide range of traits in addition to MRI based phenotypes. All of these studies included the UK biobank so cannot be considered replication cohorts. Two genes identified in this study were reported in Backman *et al.* (2021)[19] as subthreshold associations; *FLII* with ventricular volume (Volume of Lateral Ventricle right hemisphere) and *SCUBE2* with Cerebellar volume (Volume of Cerebellum Cortex left hemisphere). *STAB1* was the only gene reported as associated with a functional MRI based phenotype in Cirulli *et al.* (2020)[7], this variant was also suggestively significant in Backman *et al.* (2021)[19]. In our study, the most significant association with *STAB1* was with cuneus volume (P=0.016) and the association was barely nominal. We queried the 24 genes associated in this study with genebass (https://app.genebass.org/) to identify the associations in Karczewski *et al.* (2022)[8], we found that *PKD1*, *TNS3*, *SPSB3*, and *C17orf100* were significantly associated with a phenotype in their study ($P<6.7x10^{-7}$). However, none were associated with an MRI phenotype. *SPSB3* was associated with height in their study and total brain volume in our study, height and brain volume are correlated with each other[23] which may explain these associations. While these studies are extensive and include brain imaging phenotypes, there is little interpretation of brain volume phenotype associations in the manuscripts. The low overlap in findings between this study and previous studies is likely due to the lack of overlap between the phenotypes; these studies found results with non-volume MRI phenotypes (*STAB1* and T2* functional MRI) or did not combine different brain regions together (bilaterial vs unilateral hemispheres).

#### Gene evidence

We rated the soundness of the statistical association between the SWS and GWS genes based on evidence in previous literature. We have assigned tiers of confidence based on the results in this study and previous studies. Tier 1 represents associations that have direct support in previous literature with the same phenotype and same effect direction. Tier 2 represents associations that have convincing connections to brain volume or brain development phenotypes in previous literature. Tier 3 represents associations that have vague connections to brain related phenotypes. Tier 4 represents associations that have no connection to brain related phenotypes in previous literature. We categorised 2 gene phenotype associations as tier 1 (*PTEN*-total brain volume and *FA2H*-white matter), 5 gene phenotype associations are tier 2, 6 gene phenotype associations as tier 3, and 13 gene phenotype associations as tier 4 (**Supplementary Table 4**). The effect size estimates in this section are in units of $mm^3$ after conditioning on total brain volume with the exception of gene associations with total brain volume where the units are just $mm^3$.

##### *PTEN*

A burden of singleton variants (high confidence pLOF and missense) in *PTEN* (ENSG00000171862) was positively associated with total brain volume (GENE_P=$2.61 \times 10^{-7}$, P_burden=$8.93 \times 10^{-8}$, N_variants=6). One stop gained variant (10:87961095:C:T/rs121909231) and two missense variants (10:87925550:T:C/rs398123317 and 10:87894082:A:G/rs786204915) explained ~35%, ~23%, and ~21% of the gene association alone. The stop gained variant and one of the missense variants (p.Arg335Ter and p.Tyr68His in ENST00000371953) were labelled as pathogenic in ClinVar for multiple disorders including macrocephaly. All of these three variants were singletons with positive associations with total brain volume. 10:87961095:C:T (p.Arg335Ter) has been reported before in a child with macrocephaly[24]. Multiple mutations in *PTEN* have been linked to macrocephaly previously[25–27]. Previous literature has confirmed the role of *PTEN* and one of the variants highlighted in this study as relevant to macrocephaly which aligns with our finding of an increase in total brain volume in carriers of mutations in *PTEN*. The previous literature found the same effect of mutations in *PTEN* on total brain volume so this finding has been categorised as tier 1.

*FA2H*

A burden of variants (high confidence pLOF and missense) with MAF <0.001 in *FA2H* (ENSG00000103089) was positively associated with cortical volume (GENE_P=3.12x10$^{-12}$, P_burden=2.68x10$^{-13}$, N_variants=28) and negatively associated with white matter volume (GENE_P=1.16x10$^{-8}$, P_burden=1.10x10$^{-9}$, N_variants=28). A single high confidence missense variant (16:74774524:C:T/rs527421775) explained ~40% and ~30% of the association with the cortical grey matter and white matter respectively. The effect size of this variant was positive for cortical volume and negative for white matter volume (Beta_cortical_grey_matter=14251, SE_cortical_grey_matter=2986, P_cortical_grey_matter=1.82x10$^{-6}$, Beta_whitematter= -10568, SE_whitematter=2961, P_whitematter=3.58x10$^{-4}$, MAF=4.90x10$^{-4}$, MAC=36). This variant (p.Glu78Lys in ENST00000219368) has conflicting evidence in ClinVar for spastic paraplegia, a disease related to muscle tightness and weakness and was reported as 'unknown significance' in a study of *FA2H* related diseases[28]. Mutations in *FA2H* have been associated with white matter degeneration[10]. Previous literature supports the role of *FA2H* in white matter degeneration which matches the association found in this study so *FA2H* has be categorised as tier 1 for white matter.

No previous literature supported the role of *FA2H* in increasing cortical grey matter volume. It is possible that the cortical grey matter takes up a larger share of the total brain volume when the white matter volume is reduced leading to a positive association between *FA2H* and cortical grey matter volume. This theory is supported by the loss of association when total brain volume was removed as a covariate (P=3.12x10$^{-12}$ to P=0.045) where the association with white matter decreased but did not disappear (P=1.16x10$^{-8}$ to P=3.85x10$^{-4}$). In addition, the median white matter volume of carriers of p.Glu78Lys was significantly smaller (0.21 standard deviations) than non-carriers (t = 1.92, df = 35.06, p-value = 0.031; one-sided); whereas the cortical grey matter volume of carriers was not significantly different to non-carriers (0.063 standard deviations larger; t = -1.0953, df = 35.067, p-value = 0.1404; one-sided). This suggests that the association with cortical grey matter is a statistical artefact of the influence on white matter so the associations between *FA2H* and cortical grey matter has been categorised as tier 4.

*DISP1*

An aggregation of variants (all pLOF and missense) with MAF <0.01 in *DISP1* (ENSG00000154309) was associated with cerebellar volume (GENE_P= $1.36 \times 10^{-8}$, P_SKAT= $1.38 \times 10^{-9}$, N_variants=223). A single variant (1:223004684:T:C; rs144673025) explained ~67% of the overall gene association. This variant was a high confidence missense variant (p.Met1096Thr in ENST00000675850) with a negative association with cerebellar volume in the single variant association analysis (Beta=-2363, SE=425, P=$2.69 \times 10^{-8}$, MAC=506, MAF=0.0069). This variant was measured in a study of holoprosencephaly where carriers of this variant appeared to have normal MRI and no facial abnormalities if no other mutations were identified in the carriers[3]. This suggests that p.Met1096Thr does not cause a severe phenotype alone. Other mutations in *DISP1* have been associated with brain and facial abnormalities and are thought to influence morphogenesis through interactions with Sonic hedgehog protein[5]. One study identified that some patients with holoprosencephaly can also present with decreased cerebellar volume (cerebellar hypoplasia)[29] but no other study has reported that observation. Based on this evidence, mutations in *DISP1* can cause severe brain malformation; however, the previous literature did not directly connect mutations in *DISP1* to cerebellar volume so this finding has been categorised as tier 2.

*SCUBE2*

A burden of variants (all pLOF and missense) with MAF <0.001 in *SCUBE2* (ENSG00000175356) was negatively associated with cerebellar volume (GENE_P=$6.73 \times 10^{-8}$, P_burden=$5.47 \times 10^{-9}$, N_variants=195). Two missense variants (11:9066793:C:T/rs146308663 and 11:9047407:G:A/rs148718644) explained ~21% and ~12% of the gene association, respectively. Both variants were missense variants (p.Gly222Ser and p.Arg651Cys in ENST00000649792). Both variants were associated with reduced cerebellar volume (11:9066793:C:T: Beta= -3481, SE=1137, P=0.0022, MAF=$9.53 \times 10^{-4}$, MAC=70; 11:9047407:G:A: Beta= -4258, SE=1830, P=0.02, MAF=$3.67 \times 10^{-4}$, MAC=27). We could not find any previous literature to connect these variants to any phenotypes. *SCUBE2*, like *DISP1*, encodes a protein involved in the sonic hedgehog signalling pathway[30] and mutations in *SCUBE2* in combination with mutations in other genes can cause holoprosencephaly[4]. As with *DISP1*, mutations in SCUBE2 may not be enough to cause holoprosencephaly but could be sufficient to cause more moderate phenotypes, such as lower cerebellar volume. Mutations in *SCUBE2* have been associated with brain formation and the association of two genes in the same pathway (*DISP1* and *SCUBE2* in sonic hedgehog signalling) with the same phenotype in the same direction does add support for these findings. However, there was not any previous literature to directly support the association between mutations in *SCUBE2* and reduced cerebellar volume so this finding has been categorised as tier 2.

#### *ADAMTS18*

A burden of variants (high confidence pLOF and missense) with MAF <0.001 in *ADAMTS18* (ENSG00000140873) was positively associated with pars opercularis volume (GENE_P=2.54x$10^{-8}$, P_burden=1.94x$10^{-9}$, N_variants=87). Two high confidence missense variants (16:77362110:G:A/rs144011185 and 16:77293237:C:A/rs145095974) and 1 stop gained variant (16:77291288:C:T/rs139516327) explained ~11-18% of the gene association alone. All three of these variants had positive effect sizes and relatively low MAC (16:77362110:G:A: Beta= 281, SE=124, P=0.023, MAF=2.04x$10^{-4}$, MAC=15; 16:77293237:C:A: Beta= 396, SE=128, P=0.0020, MAF=1.90x$10^{-4}$, MAC=14; 16:77291288:C:T: Beta= 261, SE=102, P=0.011, MAF=2.99x$10^{-4}$, MAC=22). 16:77291288:C:T (p.Trp1127X in ENST00000282849) was reported as likely pathogenic in ClinVar for Microcornea-myopic chorioretinal atrophy, a developmental syndromic disorder characterised by abnormal eye development caused by mutations in *ADAMTS18*. Mutations in *ADAMTS18* appear to have wide ranging effects on multiple tissues including the brain[31] and potentially white matter integrity[32]. A study of *ADAMTS18* mouse knockouts showed higher dendritic branching complexity and spine density in hippocampal dentate gyrus[33], which may ultimately influence brain volume but it is difficult to connect this to pars opercularis volume specifically. While mutations in *ADAMTS18* may be related to brain properties and development, it is difficult to connect these specific mutations with increased pars opercularis volume specifically so this finding has been categorised as tier 2.

#### *GPATCH11*

An aggregation of variants (high confidence pLOF and missense) with MAF <0.01 in *GPATCH11* (ENSG00000152133) was associated with caudate volume (GENE_P=3.54x$10^{-7}$, P_ACATV=5.40x$10^{-8}$, N_variants=11). One stop gained variant (2:37092226:C:T/ rs200868969) explained ~95% of the gene association. This variant (p.Arg171Ter in ENST00000674370) was positively associated with caudate volume (Beta=839.97, SE=148.38, P=1.50x$10^{-8}$, MAF=6.81x$10^{-5}$, MAC=5). We were unable to find any mention of this variant in previous literature. A similar mutation upstream (p.Arg152Ter) was associated with early onset and severe retinal dystrophy with neurological impairment and facial dysmorphia in ClinVar. A recent preprint[34] describes four families with mutations in *GPATCH11* including (p.Arg152Ter), where all carriers seem to have normal MRI phenotypes, but one carrier of a splice variant was described as having macrocephaly. Our study finds an association with increased caudate volume but not total brain volume (P=0.028) which does not match the symptoms reported in previous literature. It seems that *GPATCH11* is relevant to brain development but not to the exact phenotype identified in this study so it has been categorised as tier 2.

*KMT5B*

An aggregation of variants (pLOF and missense) with MAF <0.01 in *KMT5B* (ENSG00000110066) was associated with banks of the superior temporal sulcus volume (GENE_P=8.14x$10^{-7}$, P_ ADD-BURDEN-SBAT_POS=1.03x$10^{-7}$, N_variants=97). The SBAT test could not be used for LOVO analyses but we prioritised two variants (11:68171022:A:C/rs1555027828 and 11:68180201:C:T) which explained ~60% and ~29% of the association from the most significant burden test (P=2.73x$10^{-5}$). The variant which explained the most significance was a missense variant (p.Cys324Gly in ENST00000304363) and the other was a splice acceptor variant that was labelled as benign in ClinVar. Both of these variants were singletons and positively associated with banks of the superior temporal sulcus volume. Mutations in *KMT5B* have been linked to developmental delay and macrocephaly[35] and our study mutations were linked with increased banks of the superior temporal sulcus volume; however, we did not observe an association with total brain volume (P=0.74). Previous literature supports the role of KMT5B with brain related phenotypes but not with banks of the superior temporal sulcus volume specifically, so this association has been categorised as tier 2.

*DOC2A*

An aggregation of variants (high confidence pLOF and missense) with MAF <0.01 in *DOC2A* (ENSG00000149927) was associated with white matter volume (GENE_P=6.00x$10^{-8}$, P_ACATV=1.01x$10^{-8}$, N_variants=29). One missense variant (16:30006906:C:T/rs958889356) explained ~96% of the gene association. This variant (p.Gly253Ser in ENST00000350119) was positively associated with white matter volume (Beta=49727.4, SE=7939.72, P=3.77x$10^{-10}$, MAF=6.81x$10^{-5}$, MAC=5). We were unable to find any mention of this variant in previous literature. *DOC2A* was associated with Alzheimer's disease (AD) through common variants in a study[36] which included the UKB. The lead variant in that study was a missense variant negatively associated with AD risk and the authors suggested that increased *DOC2A* expression was linked to increased AD risk. White matter changes are not defining symptoms AD but reduced white matter has been observed in AD patients[37]. Our study found that a missense variant in *DOC2A* was associated with increased white matter which aligns with Bellenguez *et al.* (2022) which found that a missense variant in *DOC2A* was negatively associated with AD risk. There is a possible connection between *DOC2A* and white matter through AD risk but the connection is tenuous and based on overlapping samples so this association has been categorised as tier 3.

##### *PRUNE2*

An aggregation of variants (all pLOF and missense) with MAF <0.01 in *PRUNE2* (ENSG00000106772) was associated with brain stem volume (GENE_P= $1.36x10^{-6}$, P_ACATV= $1.97x10^{-7}$, N_variants=488). A single missense variant (9:76711142:G:C/rs185829648, p.Leu378Val in ENST00000376718) explained 96% of the gene association. This variant was negatively associated with brain stem volume (Beta= -370, SE=66, P=$2.09x10^{-8}$, MAF=0.0073, MAC=538). We were unable to find any previous literature relating to this variant specifically. *PRUNE2* has been shown to be specifically expressed in spinal cord and brainstem in mice[38]. Two previous GWAS have found suggestive associations with Alzheimer's disease (AD)[39] and hippocampal atrophy in AD patients[40]. *PRUNE2* encodes the protein prune homolog 2, *PRUNE1*, which encodes the protein prune homolog 1, has been linked to a neurodevelopmental disorder with microcephaly and variable brain anomalies[41]. It is possible that *PRUNE1* and *PRUNE2* perform similar roles and mutations in *PRUNE2* may influence brain development as well; however, whether *PRUNE1* and *PRUNE2* perform similar roles is not clear. There is some evidence based on gene expression in mice connecting *PRUNE2* with the brainstem, and there were some previous associations with variants in *PRUNE2* with neurodegeneration in an AD GWAS using common variants. However, the connection between *PRUNE2* and decreased brainstem volume or any brain related phenotype is not clear in the literature so this finding has been categorised as tier 3.

##### *CCM2L*

An aggregation of variants (all pLOF and missense) with MAF <0.01 in *CCM2L* (ENSG00000101331) was associated with pars opercularis volume (GENE_P= $2.46x10^{-7}$, P_BURDEN-SBAT_POS= $1.33x10^{-7}$, N_variants=127). LOVO analysis did not prioritise a single variant. The most significant variant in the single variant association tests was 20:32015021:C:G (Beta= 1726, SE=479, P=$3.14x10^{-4}$, MAF=$1.36x10^{-5}$, MAC=1). This variant was a missense variant (p.Leu50Val in ENST00000452892). We could not find any previous literature about this variant. A homolog of *CCM2L* (*CCM2*) is a well-known risk gene for cerebral cavernous malformations; however, the function of *CCM2L* is not well understood[42]. Recent studies in zebrafish[43] and mice[44] suggest that *CCM2L* is relevant in the same pathways as *CCM2* and mutations can exacerbate cerebral cavernous malformations in combination with *CCM2* mutations. There is some evidence that loss of *CCM2L* can rescue the effects of *CCM2* cerebral cavernous malformations which contradicts the previously mentioned study on mice[45]. It is difficult to connect *CCM2L* to pars opercularis volume based on previous literature because patients affected by cerebral cavernous malformations seem to be affected in multiple brain regions[42] and it is not clear how *CCM2L* contributes to cavernous malformations. This finding has been categorised as tier 3.

*SPSB3*

An aggregation of variants (all pLOF and missense) with MAF <0.01 in *SPSB3* (ENSG00000162032) was associated with total brain volume (GENE_P= $3.27x10^{-7}$, P_SKAT= $2.50x10^{-8}$, N_variants=85). Two missense variants (16:1778029:G:A/rs35816944 and 16:1777132:G:A/rs147735377) explained ~59 and ~18% of the gene association alone. These variants (p.Ser171Leu and p.Arg345Trp in ENST00000566339) were negatively associated with total brain volume (16:1778029:G:A: Beta=-16011, SE=3289, P=$1.13x10^{-6}$, MAF=0.0071, MAC=516; 16:1777132:G:A: Beta=-11830, SE=3794, P=0.0018, MAF=0.0053, MAC=390). 16:1778029:G:A has been reported as significant in various studies of body measurements in the UK biobank, like height[46] and appendicular lean mass[47]. Little is known about the role of *SPSB3*; however, it has been predicted to contain a transcription binding site for Tgif1 and mutations in this protein are linked to holoprosencephaly[48]. It is possible that *SPSB3* influences brain development through interaction with Tgif1, a known homeobox protein but there is no direct evidence in previous literature to support this. This finding has been categorised as tier 3.

*EVI2A*

An aggregation of variants (all pLOF and missense) with MAF <0.01 in *EVI2A* (ENSG00000126860) was associated with total brain volume (GENE_P= $1.04x10^{-7}$, P_SKAT=$1.54x10^{-8}$, N_variants=50). A single missense variant (17:31318487:T:C/rs144778786) explains ~90% of the gene significance. This variant (p.Gln176Arg in ENST00000462804) was negatively associated with brain volume in the single variant association analysis (Beta=-18780, SE=3300, P=$1.27x10^{-8}$, MAF=0.0071, MAC=520). We were unable to find reports of this specific variant in previous literature. *EVI2A* encodes ecotropic viral integration site 2 A and appears to be relevant to multiple cancers[49] but we were unable to find previous literature connecting it to brain volume. EVI2A exists within intron 35 of *NF1*, which is transcribed from the opposite strand[50]. Loss of function and intronic mutations in *NF1* have been linked to tumours in the nervous system[51].

These mutations have also been linked with increased brain volume[52]. The variant in *EVI2A* associated with brain volume in this study was located within intron 35 of *NF1*. It has been suggested that variants within introns on *NF1* can influence disease by activation of cryptic splice sites[51], so it is possible that the variant associated with brain volume in this study could activate a cryptic splice site to cause decreased brain volume. However, this would imply that the variant identified in our study is gain of function variant in *NF1* as loss of function variants were associated with increased brain volume in previous literature. The evidence to connect this finding to total brain volume is tenuous but present so this finding has been categorised as tier 3.

*NDUFA2*

A burden of variants (high confidence pLOF and missense) with MAF <0.0001 in *NDUFA2* (ENSG00000131495) was positively associated with brainstem volume (GENE_P=1.94x$10^{-7}$, P_burden=6.75x$10^{-8}$, N_variants=6). Three variants (5:140647310:G:A, 5:140647254:A:G, and 5:140647288:G:A/rs138906500) explained ~46%, ~28%, and ~27% of the gene association. 5:140647310:G:A and 5:140647288:G:A were missense variants (p.Pro52Ser and p.Ser59Phe in ENST00000252102) and 5:140647254:A:G was a splice donor variant. All of these three variants were positively associated with brainstem volume and had a MAC of 1 or 2. A child with Leigh syndrome was identified as a carrier of a splice site variant (c.208+5G>A) close to the splice donor variant identified in this study (c.208+2T>C)[53]. This child presented with multiple symptoms of Leigh syndrome, including demyelination and subacute necrotizing encephalomyelopathy, and ultimately died before 1 year after birth. Another study of two patients found that *NDUFA2* mutations were associated with white matter abnormalities[54]. There appears to be a connection between *NDUFA2* and brain abnormalities but since *NDUFA2* is a vital part of the electron transport chain, mutations in *NDUFA2* are expected to have (and have been observed in[53,54]) a wide-ranging effect in multiple areas of the body. Additionally, it is not clear how white matter lesions could cause the association of increased brainstem volume in our study. While *NDUFA2* may be relevant to brain development, there is not much evidence to support the association of NDUFA2 with increase brainstem volume in our study. This finding has been categorised as tier 3.

*PKD1*

A burden of variants (all pLOF and missense) with MAF <0.01 in *PKD1* (ENSG00000008710) was positively associated with insula volume (GENE_P= 3.00x$10^{-9}$, P_burden= 2.16x$10^{-7}$, Beta= 31.61, N_variants=1373). LOVO analyses highlighted one missense variant (16:2090293:C:T/rs148478410) which explained ~11% of total burden association. This missense variant (p.Val4146Ile in ENST00000262304) had a positive effect size on insula volume in single variant association (16:2090293:C:T: Beta=87.88, SE=34.42, P=0.011, MAF=0.0032, MAC=234). This variant has been associated with appendicular lean mass in the UKB previously[47] (P=1.2x10$^{-6}$, Beta=0.0827). This suggest that this variant affects general body composition and may not be specific to the insula. In our study *PKD1* showed almost no association with total brain volume (P=0.042) and the association of *PKD1* with insula volume was found after including total brain volume as a covariate. Variants in *PKD1* are known to cause autosomal dominant polycystic kidney disease (not 16:2090293:C:T)[55]. There is no evidence in previous literature to connect *PKD1* with brain phenotypes so this association has been categorised as tier 4.

##### *CENPO*

An aggregation of variants (all pLOF and missense) with MAF <0.01 in *CENPO* (ENSG00000138092) was associated with white matter volume (GENE_P=1.26x10$^{-7}$, P_ACATV=1.58x10$^{-8}$, N_variants=67). One missense variant (2:24814425:A:G/ rs34788696) explained ~93% of the gene association. This variant (p.Asn89Ser in ENST00000380834) was positively associated with white matter volume (Beta=51086.8, SE=7938.32, P=1.23x10$^{-10}$, MAF=6.81x10$^{-5}$, MAC=5). We were unable to find any mention of this variant in previous literature. *CENPO* encodes a centromere protein and has been associated with multiple diseases but has not been associated with brain related phenotypes[56]. We could not find any previous literature to connect to CENPO to any brain related phenotypes so this association has been categorised as tier 4.

##### *TNS3*

An aggregation of variants (high confidence pLOF and missense) with MAF <0.01 in *TNS3* (ENSG00000136205) was associated with pericalcarine volume (GENE_P=6.04x10$^{-8}$, P_ACATV=8.88x10$^{-9}$, N_variants=67). One missense variant (7:47303013:T:A/rs77924433) explained ~96% of the gene association. This variant (p.Ile1132Phe in ENST00000311160) was positively associated with pericalcarine volume (Beta=311.97, SE=51.30, P=1.19x10$^{-9}$, MAF=6.95x10$^{-4}$, MAC=51). This variant was labelled as benign in ClinVar. *TNS3* has been suggested to be relevant for bone cell[57] and oligodendrocyte[58] differentiation. We could not find any previous literature connecting *TNS3* to brain related phenotypes so this association has been categorised at tier 4.

*ZBTB48*

A burden of singletons (high confidence pLOF and missense) in *ZBTB48* (ENSG00000204859) was positively associated with white matter volume (GENE_P=2.07x$10^{-7}$, P_burden=1.28x$10^{-8}$, N_variants=4). One frameshift variant (1:6589032:GC:G/rs746737643) explained ~75% of the gene association. This variant (p.Pro630Ter in ENST00000377674) was positively associated with white matter volume (Beta= 123276, SE= 17765.5, P=3.95x$10^{-12}$, MAF=1.36x$10^{-5}$, MAC=1). We could not find any previous literature about this variant or the missense variant which explained 13% of the gene association (p.Cys409Phe in ENST00000377674). *ZBTB48* encodes a protein involved in telomere homeostasis and has been suggested to be involved in cancer and aging[59]. We were unable to find any previous literature connecting *ZBTB48* to brain related phenotypes so this association has been categorised at tier 4.

*ZFR2*

An aggregation of variants (all pLOF and missense) with MAF <0.01 in *ZFR2* (ENSG00000105278) was positively associated with white matter volume (GENE_P=1.96x$10^{-8}$, P_ACATV=2.45x$10^{-9}$, N_variants=277). One missense variant (19:3819069:C:T/ rs772588719) explained 98% of the gene association. This variant (p.Arg636Gln in ENST00000262961) was positively associated with white matter volume (Beta= 49532 SE= 7239.32, P=7.79x$10^{-12}$, MAF=8.17x$10^{-5}$, MAC=6). We could not find any previous literature about this variant. We could not find any literature to connect *ZFR2* to any brain related phenotypes so this association has been categorised at tier 4.

*RPS21*

A single singleton (high confidence missense) in *RPS21* (ENSG00000171858) was positively associated with white matter and cortical grey matter volume (White matter: GENE_P=1.94x$10^{-11}$, P_burden=1.40x$10^{-12}$, N_variants=1; Cortical grey matter: GENE_P=2.36x$10^{-6}$, P_burden=1.89x$10^{-7}$, N_variants=1). This variant (20:62387883:C:T/rs1987795982; p.Thr52Ile in ENST00000343986) was positively associated with both phenotypes in the single variant analyses. We were unable to find previous studies reporting this variant. *RPS21* encodes a protein relevant for ribosome biogenesis, cell growth, and apoptosis[60], we were unable to find previous studies supporting the role of *RPS21* in brain phenotypes. Due to the low evidence in the literature and that the association in this study was driven by one singleton, it is likely that this finding is a false positive. This finding has been categorised as tier 4 for both cortical grey matter and white matter.

*POLD4*

A single singleton (splice acceptor) in *POLD4* (ENSG00000175482) was positively associated with cortical grey matter volume (GENE_P=6.14x$10^{-10}$, P_burden=3.94x$10^{-11}$, N_variants=1). This variant (11:67353079:T:C) was positively associated with cortical grey matter volume in the single variant analyses (Beta=118231, SE=17897, P=3.94x$10^{-11}$, MAF=1.36x$10^{-5}$, MAC=1). We were unable to find previous studies reporting this variant. *POLD4* encodes DNA polymerase delta subunit 4 and overexpression has been associated with increased risk of gliomas[61]. Our study found an association between a splice acceptor in *POLD4* and an increase in cortical grey matter volume, it is a possible that this splice acceptor could lead to increase expression which could cause gliomas causing an increase in cortical grey matter volume. However, it is unknown how this splice acceptor affects *POLD4* and whether gliomas would cause an increase in cortical grey matter volume specifically. Additionally, the association was due to singleton so it is more likely that this finding was a false positive. This finding has been categorised as tier 4.

*SPMIP1*

A burden of singleton variants (high confidence pLOF and missense) in *SPMIP1* (ENSG00000272899) was positively associated with rostral anterior cingulate volume (GENE_P=1.77x$10^{-6}$, P_burden=1.65x$10^{-7}$, N_variants=3). These three variants (7:128866695:C:T/rs564757432, 7:128866453:G:A/rs1446913994, and 7:128866783:GA:G) explained ~47%, ~27%, and ~13% alone. Two of these variants (7:128866695:C:T and 7:128866453:G:A) were stop gained variants (p.Arg96Ter and p.Trp15Ter in ENST00000609480) and one (7:128866783:GA:G) was a frameshift variant (p.Tyr126ThrfsTer19 in ENST00000609480). All of these variants were singletons and had positive associations with rostral anterior cingulate volume. We were unable to find any previous literature around *SPMIP1* or the variants we identified. The association of this gene in our study was due to 3 singletons and we were unable to find previous literature connecting this gene to any phenotypes so we predict that this finding is likely a false positive. This finding has been categorised as tier 4.

*C17orf100*

A burden of variants (all confidence pLOF and missense) with MAF <0.0001 in *C17orf100* (ENSG00000256806) was positively associated with white matter volume (GENE_P=1.27x$10^{-7}$, P_burden=1.55x$10^{-8}$, N_variants=2). A frameshift variant (17:6651923:GC:G) explained ~95% of the gene association. This variant (p.Arg5GlufsTer86 in ENST00000542475) was positively associated with white matter volume (Beta=127388, SE=17752, P=7.18x$10^{-13}$, MAF=1.36x$10^{-5}$, MAC=1). We were unable to find previous literature about this variant or any previous literature linking *C17orf100* to brain related phenotypes. Considering the lack of support from previous literature and that this association was driven by one singleton, it is likely that this association is a false positive. This finding has been categorised as tier 4.

*ADCY6*

A burden of variants (high confidence pLOF and missense) with MAF <0.0001 in *ADCY6* (ENSG00000174233) was negatively associated with hippocampus volume (GENE_P=5.21x$10^{-7}$, P_burden=1.60x$10^{-8}$, N_variants=78). Three high confidence missense variants explained approximately ~13-15% of the gene association each (12:48776334:G:A/rs201227124, 12:48776301:C:T/rs758588591, 12:48776513:C:T/rs781510708). All of these variants (p.Arg518Trp, p.Glu529Lys, and p.Val484Ile in ENST00000357869) were negatively associated with hippocampal volume with a minor allele count of 4-5. We were unable to find previous literature on these specific variants. The role *ADCY6* in the human body is not well understood but a specific mutation in this gene has been linked to diseases of the peripheral nervous system[62]; however there is no evidence to support the role of *ADCY6* in brain related phenotypes. This finding has been categorised as tier 4.

*RPUSD1*

An aggregation of variants (all pLOF and missense) with MAF <0.01 in *RPUSD1* (ENSG00000007376) was associated with precuneus volume (GENE_P= 1.78x$10^{-6}$, P_SKAT= 2.07x$10^{-7}$, N_variants=112). Two missense variants (16:786149:A:G/rs3751672 and 16:786081:C:A/rs61745562) explained ~18-20% of the gene association individually. These variants (p.Leu247Pro and p.Gly270Cys in ENST00000007264) were positively associated with precuneus volume with relatively large minor allele counts 112-119. Neither of these variants have been reported in previous literature. We were also unable to find any previous literature outlining the role of *RPUSD1* in disease or brain related phenotypes. This finding has been categorised as tier 4.
